# The prognostic, predictive and clinicopathological impact of KRT81 / HNF1A- and GATA6- based transcriptional subtyping in pancreatic cancer

**DOI:** 10.1101/2024.04.29.24306532

**Authors:** Michael Guenther, Sai Agash Surendran, Volker Heinemann, Michael Haas, Stefan Boeck, Steffen Ormanns

## Abstract

**BACKGROUND:** Transcriptional subtypes of pancreatic ductal adenocarcinoma (PDAC) have prognostic implications and potential predictive functions. This study aimed to determine their clinicopathological impact in large cohorts of advanced and resected PDAC and their evolution during disease progression.

**METHODS:** The clinicopathological and prognostic implications of transcriptional subtypes determined by the expression of KRT81, HNF1A and GATA6 were examined using immunohistochemistry in advanced (n=139) and resected (n=411) PDAC samples as well as in 57 matched primary tumors and corresponding metastases. RNAseq data of 316 resected PDAC patients was analyzed for validation.

**RESULTS:** Both subtyping systems were highly interrelated. Subtypes switched during disease progression in up to 31.6% of patients. Transcriptional subtyping had a modest prognostic impact in both unstratified cohorts, but strongly improved outcomes in patients with KRT81 positive / GATA6 negative tumors treated with palliative or adjuvant gemcitabine-based chemotherapy. RNAseq expression data confirmed the findings.

**CONCLUSIONS:** Transcriptional subtypes have differential responses on palliative and adjuvant gemcitabine- based chemotherapy, but they may change during disease progression. Both employed subtyping systems are equivalent and can be used to inform clinical therapy decisions.

**CLINICAL TRIAL REGISTRY:** The clinical trial registry identifier is NCT00440167.

## INTRODUCTION

Although modest improvements in the prognosis of pancreatic ductal adenocarcinoma (PDAC) patients were achieved in the past years, mainly through innovative randomized control trials (RCT) on novel chemotherapy (CTX) regimens, the survival probability of the average PDAC patients remains desperately low ^1^, even for patients in early disease stages^2^. For the minority of patients diagnosed with resectable disease, adjuvant therapy has become standard of care, with the active but toxic regimen FOLFIRINOX as 1^st^ choice for the clinically fit patient and the less active but less toxic gemcitabine for the less fit^3^. Large sequencing studies have revealed the molecular background of the disease on the genetic as well as the transcriptional level, which resulted in the identification of transcriptional subtypes, based on the expression of specific hallmark genes^4^. Thus, several transcriptional subtyping systems were proposed, each of which showed a differential prognostic impact of the subtypes identified^4^. Some studies even proposed prognostic implications of transcriptional subtyping, which largely relied on complex, RNA-based methodologies such as RNA-sequencing (RNAseq), which are hard to establish in routine diagnostics. Therefore, efforts were made to break down complex subtyping systems to simpler approaches applicable in routine clinical practice by detecting the expression of each subtypeś hallmark genes through immunohistochemistry (IHC)^5,6^. Although these approaches resulted in valuable information on the potential clinical implications of the subtypes identified, their meaning to inform clinical decision making was modest at best^6,7^. Moreover, most studies did not examine the clinical significance of transcriptional subtypes with respect to clinicopathological parameters such as disease stage or the therapies applied -specifically in the adjuvant setting- and no study to date compared the different subtyping systems with each other in the same set of samples. Thus, in the present study, we examined the prognostic and potentially predictive impact of the two most employed IHC-based subtyping systems, i.e. the expression of KRT81 and HNF1A or GATA6 respectively, in large cohorts of advanced PDAC and resected PDAC patients, compared both subtyping systems and determined how the metastatic process may affect the predominant subtype.

## MATERIALS and METHODS

The study cohorts and the inclusion and exclusion criteria were described previously^8^. Archival formalin-fixed paraffin embedded (FFPE) histologically confirmed tumor tissue of primary tumors and metastatic tissue was collected from the pathology laboratories where the diagnosis of PDAC was first established. Patients’ overall survival (OS), disease free survival (DFS) as well as progression free survival (PFS) was calculated as described before^8^. Written informed consent for the use of tumor material and clinical data was obtained from advanced PDAC patients upon study enrollment or before palliative chemotherapy initiation. The ethics committee of the medical faculty LMU approved the use of patient material and data in the resected PDAC cohort (project 20-081). Tissue microarray (TMA) construction was described before^8^. GATA6 expression was detected on four µm thick sections by immunohistochemistry using an anti-GATA6 polyclonal rabbit antibody (PA1-104, Thermo Fisher, Germering, Germany) at a 1:200 dilution. Immunohistochemical detection of KRT81 and HNF1A was performed as described previously^9^. Appropriate positive controls were included in each staining run (human normal tonsil for KRT81, duodenal mucosa for HNF1A and normal exocrine pancreas for GATA6, suppl. figure S1 A - C). The expression pattern and expression strength were independently evaluated by two pathologists (MG, SO) blinded to the patient outcome and discrepant cases were discussed until agreement was reached. Tumors were classified as follows: samples with ≥ 30% KRT81- or HNF1A-positive tumor cells were considered positive for each marker. For GATA6 expression, tumors with distinct nuclear staining were considered GATA6 positive. Microphotographs were acquired as described previously^8^. Kaplan-Meier curves, Cox regression analyses and cross tabulations were computed using SPSS software (IBM, Ehningen, Germany), considering a p-value of ≤ 0.05 as statistically significant. Propensity score matching, as well as the analysis of publicly available gene expression data and the patientś corresponding clinical data was carried out as described before ^8^. RNA sequencing data and clinical information of the ICGC-CA cohort were downloaded from International Cancer Genome Consortium (ICGC) data portal ^10^. Genomic analysis and data visualization was conducted using cBioPortal for Cancer Genomics^11^.

## RESULTS

### Transcriptional subtypes according to the expression of KRT81, HNF1A and GATA6 widely overlap but may change during metastatic progression

Of the 139 samples in the advanced PDAC (aPDAC) cohort, 36.0% (n=50) were KRT81+, 36.0% (n=50) were double negative and 28.0% (n=39) were HNF1A+, whereas 60.4% (n=84) were GATA6- and 39.6% (n=55) were positive for GATA6. In the resected PDAC (rPDAC) cohort, 39.9% (n=164) were KRT81+, 43.6% (n=179) were double negative and 16.5% (n=68) were HNFA1+, whereas 41.1% (n=169) displayed GATA6 expression and 58.9% (n=242) were GATA6- (figure 1A). Tumors displaying expression of both KRT81 and HNF1A (double positive), which was the case in 16.5% of the samples in the aPDAC cohort and 7.1% in the rPDAC cohort, were categorized according to the marker showing a predominant expression pattern. This approach was backed by highly similar prognostic implications of the thus assessed subtypes in the double positive cases compared to the single positive ones (suppl. table S1). Both subtyping systems overlapped widely in both cohorts (χ^2^ p<0.001 each), with the HNF1A+ subtype being mostly GATA6+ (suppl. Table S2). However, a significant proportion of KRT81+ samples displayed GATA6 expression in both cohorts and no clear trend towards a GATA6-based subtype was observed for the double negative samples, which were rather GATA6- in the aPDAC cohort and rather GATA6+ in the rPDAC cohort (figure 1B and C). For both systems, the assessment of each tumors subtype remained remarkably stable when several tumor tissue punches from different FFPE blocks were compared separately (suppl. table S3).

**Figure 1.**
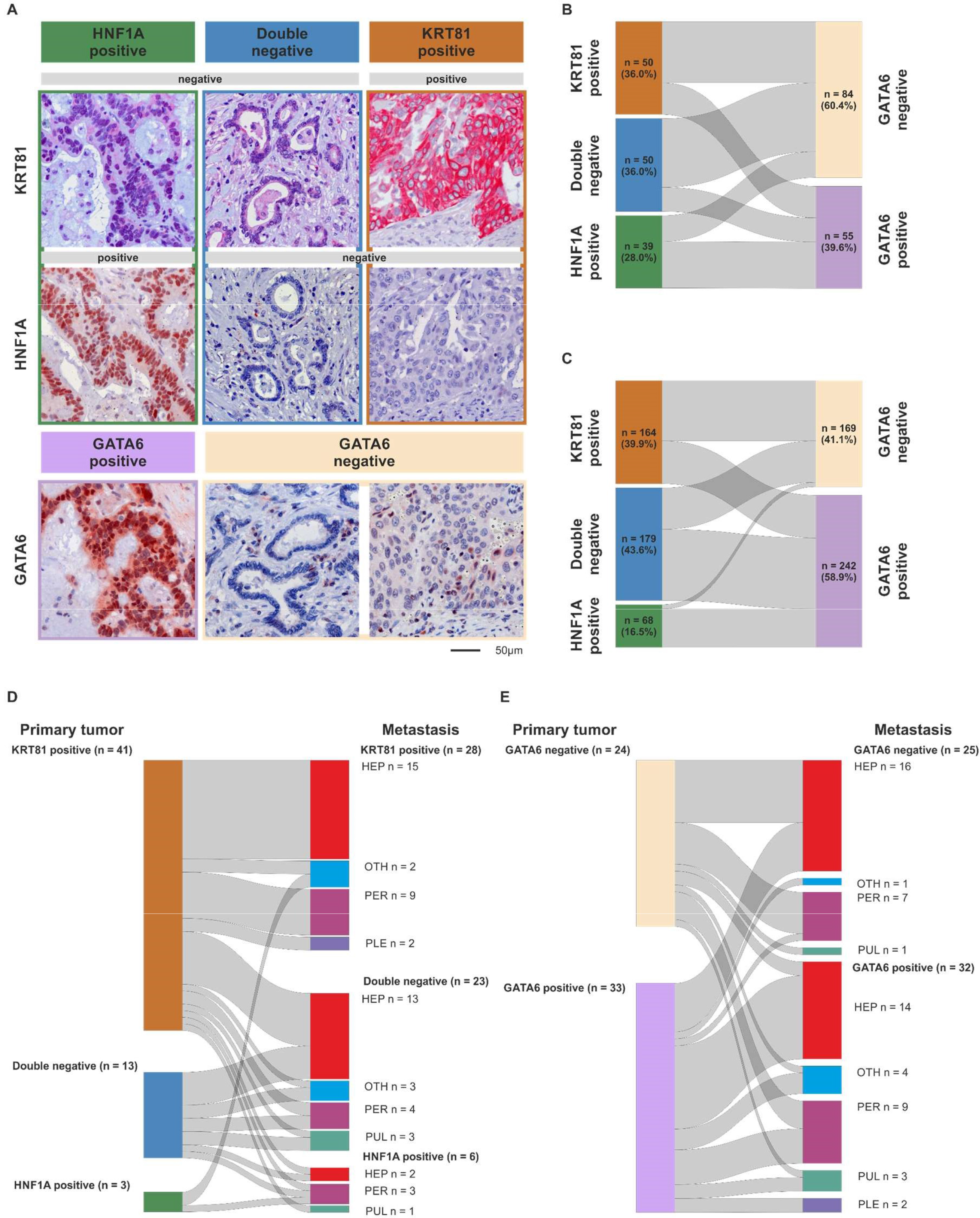
Differential expression of KRT81, HNF1A and GATA6 in pancreatic cancer Immunohistochemical detection *of KRT81, HNF1A and GATA6* expression in exemplary PDAC samples. 200-fold magnification. Scale bar indicates 50 µm (A). Comparison of the tumorś transcriptional subtypes based on the expression of KRT81 / HNF1A or GATA6 in advanced (B) and resected PDAC (C) as well as between the primary tumor and its corresponding metastasis using subtyping based on KRT81 / HNF1A-expression (D) or GATA6-expression (E).

To assess how metastatic progression may affect the tumors subtype, we examined to which extent the primary tumorś and the metastatic tissueś subtypes were interrelated in synchronous and metachronous metastases to its corresponding primary in 57 cases. There was a strong correlation between the primarieś and the metastaseś subtype (p=0.001), but we observed a subtype switch according to KRT81 / HNF1A expression in 18 cases (31.6%). This was not associated to synchronous or metachronous metastasis (suppl. table S4) but came with a highly significant trend towards a prognostically more favorable subtype (suppl. Table S5). More than half (n=28) of the n=41 KRT81+ cases remained KRT81+ in their corresponding metastasis, the rest was mostly double negative and only three cases switched to HNF1A+. Double negative cases mostly remained double negative during metastasis, whereas the few (n=3) HNF1A+ cases split between KRT81 positivity and HNF1A positivity in their metastatic tissue (figure 1 D, suppl. table S6). In contrast, the transcriptional subtype according to GATA6 expression remained stable in most cases with 9 of 34 cases switching from GATA6- to GATA6+ and 7 of 23 cases switching from GATA6+ to GATA6- (figure 1 E, suppl. table S6). Here, subtype switching occurred more frequently in synchronous metastases (suppl. table S4). Neither the primary tumorś nor the metastases subtype or the occurrence of a subtype switch was associated with metastasis localization (suppl. table S7).

### Transcriptional subtypes affect patient outcome dependent on palliative therapy in advanced PDAC patients

The baseline characteristics, outcome and clinicopathological variables of the aPDAC patient cohort were previously described^8^. In the unstratified cohort, transcriptional subtypes according to KRT81/HNF1A expression implied a statistically non-significant trend for patient OS, with HNF1A+ cases having the best, double negative cases an intermediate and KRT81+ the worst outcome (OS 6.8 vs. 9.1 vs. 10.7 months, p= 0.08, HR=1.27 95%, CI 1.02 – 1.59, suppl. figure S2 A), which was not reflected in PFS times (PFS 3.6 vs. 4.1 vs. 6.4 months, p= 0.42, HR=1.16 95% CI 0.91 – 1.48, suppl. figure S2 B). Similarly, subtyping based on GATA6 expression had no prognostic impact in the unstratified patient cohort (PFS 4.2 vs 4.3 months, p= 0.43, HR 1.16 95%CI 0.80 – 1.69; OS 7.8 vs. 9.2 months, p=0.76, HR 1.06, 95% CI 0.74 – 1.51, suppl. figure S2 C, D). As expected, only KRT81/ HNF1A based subtypes showed a trend towards statistical significance in multivariate analyses (p=0.07, suppl. table S8) and transcriptional subtypes were not associated with the patientś clinicopathological variables (table 1). Stratification of patient subgroups according to the applied type of 1^st^ line palliative treatment, revealed a significant impact of the tumors transcriptional subtype on patient outcome, as patients with double negative tumors derived the most benefit from palliative gemcitabine-based chemotherapy (pGC) compared to palliative non-gemcitabine-based chemotherapy (pnGC, PFS 6.3 vs 2.4 months, p<0.001, HR 0.26, 95%CI 0.13 – 0.52; OS 9.6 vs 5.7 months, p=0.04, HR 0.53, 95%CI 0.29 – 0.97, figure 2 A,B). Similarly, patients with KRT81+ tumors showed favorable PFS times with pGC compared to pnGC (PFS 4.5 vs. 2.2 months, p=0.02, HR 0.43, 95%CI 0.21 – 0.90 figure 2 A), but no significant impact of palliative chemotherapy on OS times (OS 6.8 vs 4.7 months, p=0.47, HR 0.78 95%CI 0.40 – 1.52, figure 2 B). Interestingly, the type of palliative chemotherapy had no significant prognostic impact in patients with HNF1A+ tumors (pnGC vs. pGC, PFS 6.6 vs. 2.7 months, p=0.28, HR 0.65 95%CI 0.30 – 1.42; OS 13.2 vs. 9.3 months, p=0.60, HR 0.82, 95%CI 0.38 – 1.75 figure 2 A, B), which was also confirmed in multivariate analyses (suppl. table S9). Applying the GATA6-based subtypes paralleled these findings, as patients with GATA6- tumors profited from pGC compared to pnGC, whereas no significant prognostic impact of the type of palliative chemotherapy was detected for patients with GATA6+ tumors (GATA6- PFS 9.3 vs 2.4 months, p<0.001, HR 0.27, 95%CI 0.15 – 0.48; OS 9.7 vs 7.2 months, p=0.04, HR 0.60, 95%CI 0.38 – 0.97; GATA6+ 5.0 vs 2.7 months, p=0.21, HR 0.65 95%CI 0.33 – 1.29; OS 7.8 vs 4.9 months, p=0.34, HR 0.72 95%CI 0.37 – 1.42, figure 2 C,D), which was confirmed for PFS in multivariate analyses (suppl. table S9).

**Figure 2.**
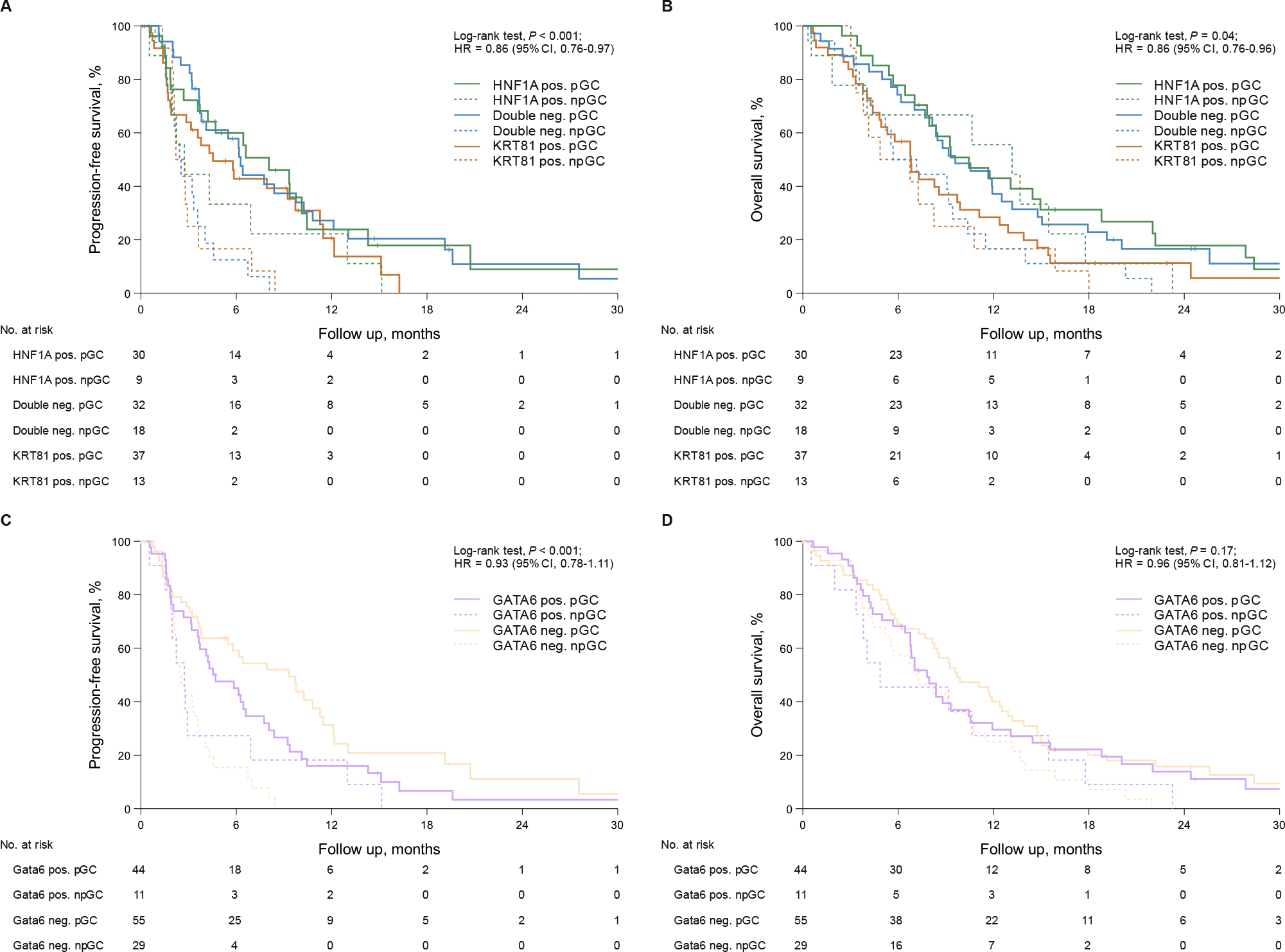
Transcriptional subtypes are associated with therapy response in 1st line gemcitabine treated advanced pancreatic cancer patients Univariate analyses (Kaplan–Meier curves and log-rank tests) for PFS and OS in the subtypes based on KRT81 / HNF1A - expression stratified by 1st line chemotherapy (A, B) as well as in the subtypes based on GATA6 - expression stratified by 1st line chemotherapy (C, D). Crossed lines indicate censored cases.

**Table 1.**
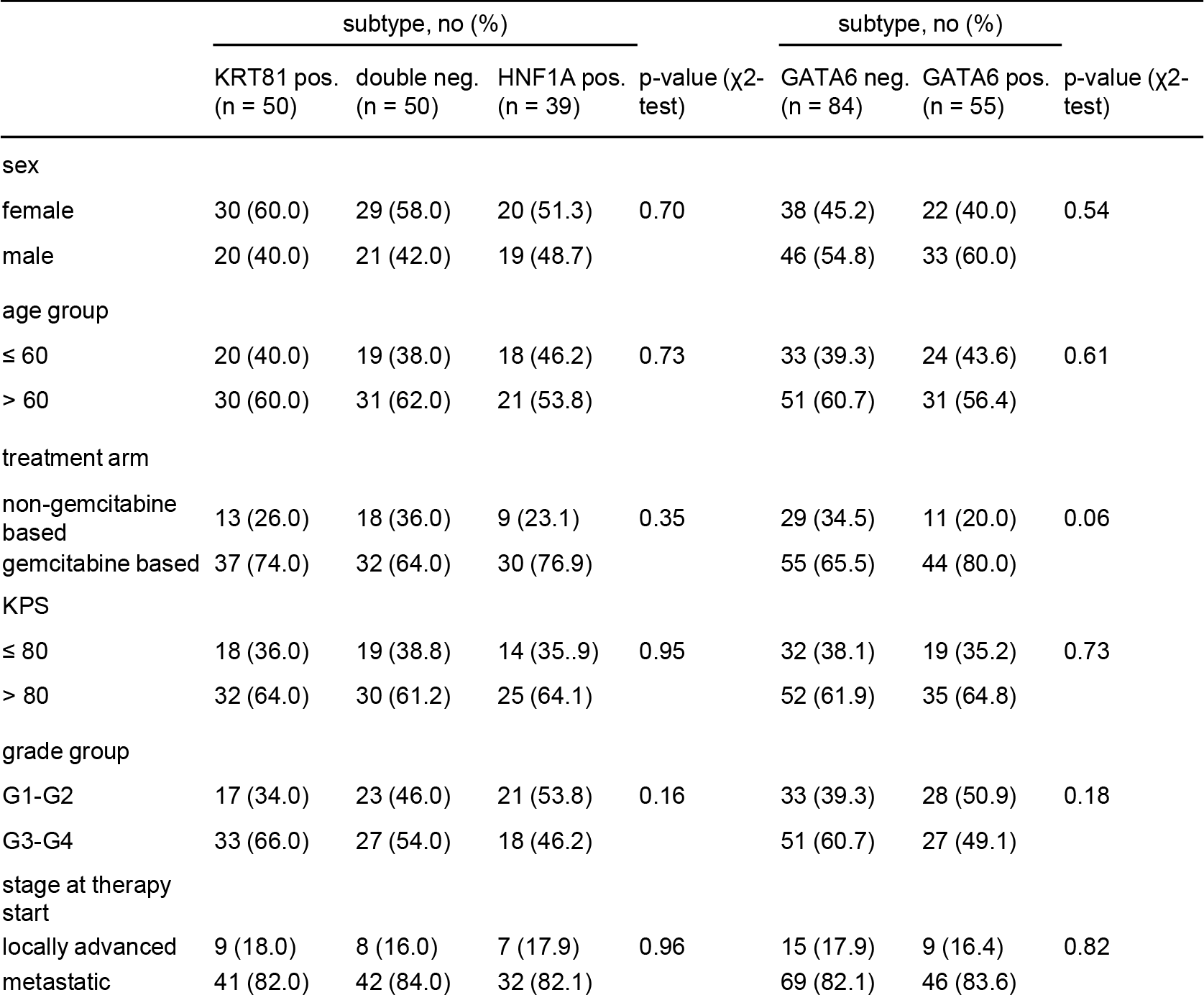
Clinicopathological patient characteristics according to KRT81 / HNF1A - expression and GATA6 - expression in the aPDAC cohort.

### Transcriptional subtypes affect patient outcome dependent on adjuvant therapy in resected PDAC patients

In the unstratified rPDAC cohort, there was a statistically non-significant impact of the tumorś subtype on outcome, with KRT81 positivity conferring the worst, double negative an intermediate and HNF1A positivity the best outcome (OS 17.1 vs. 19.3 vs. 22.3 months, p=0.34, HR 0.87, 95% CI 0.81 – 0.94; DFS 9.7 vs. 12.1 vs. 12.4 months, p= 0.47, HR 1.08, 95%CI:0.90 – 1.28, suppl. figure S3 A, B), which turned out as independent prognostic marker for OS in multivariate analyses (suppl. table S 10). Similarly, GATA6 expression was associated with a statistically not significant trend towards better prognosis (OS 20.7 vs. 15.2 months, p=0.20, HR 0.87, 95% CI 0.69 – 1.08, suppl. figure S3 C) but showed no tendency for differences in DFS (suppl. figure S3 D) although it turned out as independent prognosticator for OS in multivariate analyses (p=0.006, HR 0.73, 95%CI 0.58 – 0.91, suppl. table S10). Transcriptional subtypes were not associated to the patientśclinicopathological parameters (table 2). To test the impact of subtyping on response to adjuvant chemotherapy we calculated DFS and OS times in each subtype according to the application of adjuvant gemcitabine-based chemotherapy (aGC) compared to no or non-gemcitabine-based adjuvant chemotherapy (naGC). Interestingly, the prognostically worst KRT81+ subtype, had the strongest prognostic impact of adjuvant gemcitabine-based chemotherapy (DFS 5.0 vs. 13.7 months, p<0.001, HR 0.37, 95% CI 0.25 – 0.55; OS 8.3 vs. 31.6 months, p<0.001, HR 0.26 95%CI 0.18 – 0.37), whereas this was not the case for the double negative subtype (DFS 10.1 vs. 13.5 months, p= 0.52, HR 0.88, 95%CI 0.59 – 1.32; OS 17.7 vs. 21.0 months, p=0.08, HR 0.76, 95%CI 0.54 – 1.07) or the HNF1A+ subtype (DFS 12.8 vs. 9.5 months, p=0.39, HR 1.32, 95%CI 0.70 – 2.49; OS 20.4 vs. 23.4 months, p=0.38, HR 0.78, 95%CI 0.45 – 1.36, figure 3 A,B). Similar results were obtained for GATA6-based subtyping, where the GATA6- subtype profited significantly from adjuvant gemcitabine-based chemotherapy (DFS 4.8 vs. 15.1 months, p<0.001, HR 0.30, 95%CI 0.19 – 0.46; OS 7.0 vs. 32.2 months, p<0.001, HR 0.21, 95%CI 0.66 – 1.18), whereas it was not beneficial in the GATA6+ subtype (DFS 10.5 vs. 10.0 months, p=0.62, HR 1.09, 95%CI 0.78 – 1.51; OS 18.9 vs. 21.5 months, p= 0.42, HR 0.87, 95%CI 0.66 – 1.18, figure 3 C,D). These findings were also reflected in 5- year-survival rates (suppl. table S11). We confirmed these findings in multivariate analyses which also demonstrated the prognostic impact of the patientś clinicopathological characteristics in each subtype. Interestingly, the R-status was most important for DFS and OS in the KRT81+ and GATA6- subtypes, whereas in the HNF1A+ and the GATA6+ subtypes pN-stage had the strongest prognostic influence (suppl. table S11).

**Figure 3.**
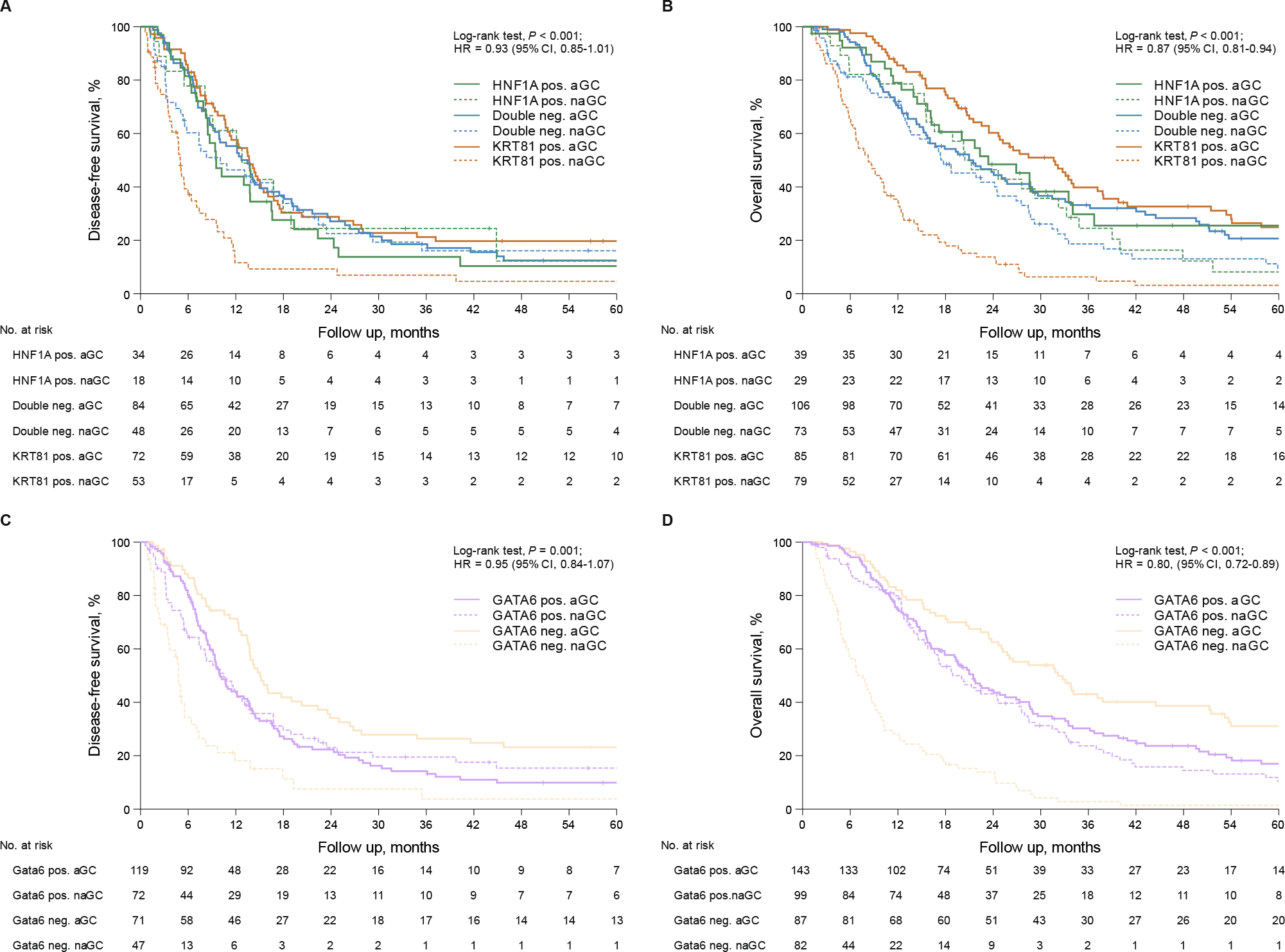
Transcriptional subtypes are associated with therapy response in resected pancreatic cancer patients treated with adjuvant gemcitabine-based chemotherapy Univariate analyses (Kaplan–Meier curves and log-rank tests) for DFS and OS in the subtypes based on KRT81 / HNF1A - expression stratified by gemcitabine-based adjuvant treatment (A, B) and in the subtypes based on GATA6 - expression stratified by gemcitabine- based adjuvant treatment (C, D). Crossed lines indicate censored cases.

**Table 2.** Clinicopathological patient characteristics according to KRT81 / HNF1A - expression and GATA6 - expression in the rPDAC cohort.

|  | subtype, no (%) |  |  | P value (χ<sup>2</sup> ) | subtype, no (%) |  | P value (χ<sup>2</sup> ) |
| --- | --- | --- | --- | --- | --- | --- | --- |
|  | KRT81-positive<br>(n=164) | Double-negative<br>(n=179) | HNF1A-positive<br>(n=68) |  | GATA6 neg.(n=169) | GATA6 pos.<br>(n=242) |  |
| sex |  |  |  |  |  |  |  |
| female | 72 (43.9) | 99 (55.3) | 29 (42.6) | 0.06 | 72 (42.6) | 128 (52.9) | 0.04 |
| male | 92 (56.1) | 80 (44.7) | 39 (57.4) |  | 97 (57.4) | 114 (47.1) |  |
| age group |  |  |  |  |  |  |  |
| ≤ 68 | 91 (55.5) | 93 (52.0) | 29 (42.6) | 0.20 | 73 (43.2) | 125 (51.7) | 0.09 |
| > 68 | 73 (44.5) | 86 (48.0) | 39 (57.4) |  | 96 (56.8) | 117 (48.3) |  |
| treatment arm, no (%) |  |  |  |  |  |  |  |
| non-gemcitabinebased | 79 (48.2) | 73 (40.8) | 29 (42.6) | 0.38 | 82 (48.5) | 99 (40.9) | 0.13 |
| gemcitabinebased | 85 (51.8) | 106 (59.2) | 39 (57.4) |  | 87 (51.5) | 143 (59.1) |  |
| UICC stage (2017) |  |  |  |  |  |  |  |
| stage IA | 6 (3.7) | 12 (6.7) | 6 (8.8) | 0.37 | 8 (4.7) | 16 (6.6) | 0.92 |
| stage IB | 33 (20.1) | 32 (17.9) | 16 (23.5) |  | 33 (19.5) | 48 (19.8) |  |
| stage IIA | 16 (9.8) | 14 (7.8) | 5 (7.4) |  | 16 (9.5) | 19 (7.9) |  |
| stage IIB | 64 (39.0) | 54 (30.2) | 18 (26.5) |  | 56 (33.1) | 80 (33.1) |  |
| stage III | 25 (15.2) | 41 (22.9) | 16 (23.5) |  | 32 (18.9) | 50 (20.7) |  |
| stage IV | 20 (12.2) | 26 (14.5) | 7 (10.3) |  | 24 (14.2) | 29 (12.0) |  |
| pT (2017) |  |  |  |  |  |  |  |
| pT1a | 1 (0.6) | 0 (0.0) | 2 (2.9) | 0.06 | 0 (0.0) | 3 (1.2) | 0.36 |
| pT1b | 1 (0.6) | 4 (2.2) | 0 (0.0) |  | 2 (1.2) | 3 (1.2) |  |
| pT1c | 11 (6.7) | 26 (14.5) | 8 (11.8) |  | 14 (8.3) | 31 (12.8) |  |
| pT2 | 104 (63.4) | 99 (55.3) | 40 (58.8) |  | 106 (62.7) | 137 (56.6) |  |
| pT3 | 46 (28.0) | 46 (25.7) | 18 (26.5) |  | 46 (27.2) | 64 (26.4) |  |
| pT4 | 1 (0.6) | 4 (2.2) | 0 (0.0) |  | 1 (0.6) | 4 (1.7) |  |
| pN (2017) |  |  |  |  |  |  |  |
| pN0 | 62 (37.8) | 71 (39.7) | 31 (45.6) | 0.18 | 69 (40.8) | 95 (39.3) | 0.88 |
| pN1 | 68 (41.5) | 62 (34.6) | 17 (25.0) |  | 61 (36.1) | 86 (35.5) |  |
| pN2 | 34 (20.7) | 46 (25.7) | 20 (29.4) |  | 39 (23.1) | 61 (25.2) |  |
| R-status |  |  |  |  |  |  |  |
| 0 | 106 (64.6) | 121 (67.7) | 48 (70.6) | 0.66 | 108 (63.9) | 167 (69.0) | 0.28 |
| 1 | 58 (35.4) | 58 (32.4) | 20 (29.4) |  | 61 (36.1) | 75 (31.0) |  |
| grade group |  |  |  |  |  |  |  |
| G1-G2 | 45 (27.4) | 53 (29.6) | 23 (33.8) | 0.62 | 51 (30.2) | 70 (28.9) | 0.78 |
| G3-G4 | 119 (72.6) | 126 (70.4) | 45 (66.2) |  | 118 (69.8) | 172 (71.1) |  |

We validated these findings in two independent publicly available PDAC cohorts (n=138 and n=178) with corresponding RNAseq-based gene expression data. In both datasets, just as in our cohort, patients with KRT81+ or GATA6- tumors derived the most benefit from gemcitabine-based adjuvant chemotherapy, whereas in the other subtypes the impact of aGC was minor (suppl. figure S4).

To explore a potential mechanistic background for the observed gemcitabine-resistance in the patients with HNF1A+/ GATA6+ tumors, we tested the association of the expression of HNF1A, KRT81 and GATA6 with the expression of gemcitabine-resistance associated genes in the same RNAseq-based gene expression datasets. In both cohorts, there was a strong positive correlation with HNF1A and GATA6 expression and an inverse correlation for KRT81 with the expression of *ABCC3* and *MVP*, both known to cause gemcitabine-resistance *in vitro* and *in vivo*^12,13^ (suppl. figure S5). Of note, after adjusting for multiple testing, transcriptional subtypes did not correlate with specific molecular alterations on the genomic level (suppl. figure S6).

## DISCUSSION

Transcriptional subtyping in pancreatic cancer using different systems of hallmark genes and detection techniques and its potential clinical or biological impact has been reported several times to date ^5–7,9,14–25^ (table 3). There is a certain consensus that there are at least two subtypes, mostly termed “basal” or “quasi-mesenchymal” and “classical”^26^. Whereas the first one is often associated with poorer outcome and sometimes higher tumor grade, exemplified by the adenosquamous subtype of PDAC, the latter is associated with better prognosis and better therapy response to FOLFIRINOX (FFX)^14,27^. Other previously proposed transcriptional subtypes, such as the so called “immunogenic” or “ADEX” subtype by Bailey et al (QUOTE), could not be reproduced entirely by other groups ^28^. Thus, their existence remains a matter of debate to date. None of the previously published studies on transcriptional subtyping in PDAC examined their predictive impact on currently still widely employed therapies. Moreover, none of the IHC-based subtyping systems were compared in the same set of patient samples as yet and a potential subtype switch during metastatic progression -which eventually occurs in the majority of PDAC patients- has not been examined either. In this study, we employed and compared the two most widespread subtyping systems based on the robust immunohistochemical detection of KRT81, HNF1A or GATA6. We show that both systems overlap widely and can be employed equally, which offers a cost efficient approach to personalized cancer therapy as specific subtypes respond differentially to therapy. For instance, patients with KRT81+ / GATA6- tumors profit better from gemcitabine-based chemotherapy in the palliative but also the adjuvant setting compared to their KRT81-/ GATA+ counterparts in our cohorts, which is backed by previously published data ^6,29^. Thus, transcriptional subtyping by IHC could be used to inform therapy decisions in the routine clinical setting, even in cases with scarce or low-quality tumor material in which RNA-based approaches tend to fail. Our approach to categorize according to the most dominant subtype in the KRT81/HNF1A system, resolved the issue of double positive cases, which were excluded from analysis in previously published studies ^5,7,22,23^. Our novel finding that transcriptional subtypes may switch during metastatic progression may explain differential therapy responses of primary tumors and metastases and implies a rationale to re-biopsy metastatic tissue to inform a subtype-based therapy choice. We also show that the R-status has a much stronger prognostic impact in KRT81+ / GATA6- tumors. Thus, in this subtype, it is much more important to achieve a wide R0-resection compared to others, which should be considered during resection and may justify a more aggressive surgical approach.

**Table 3.**
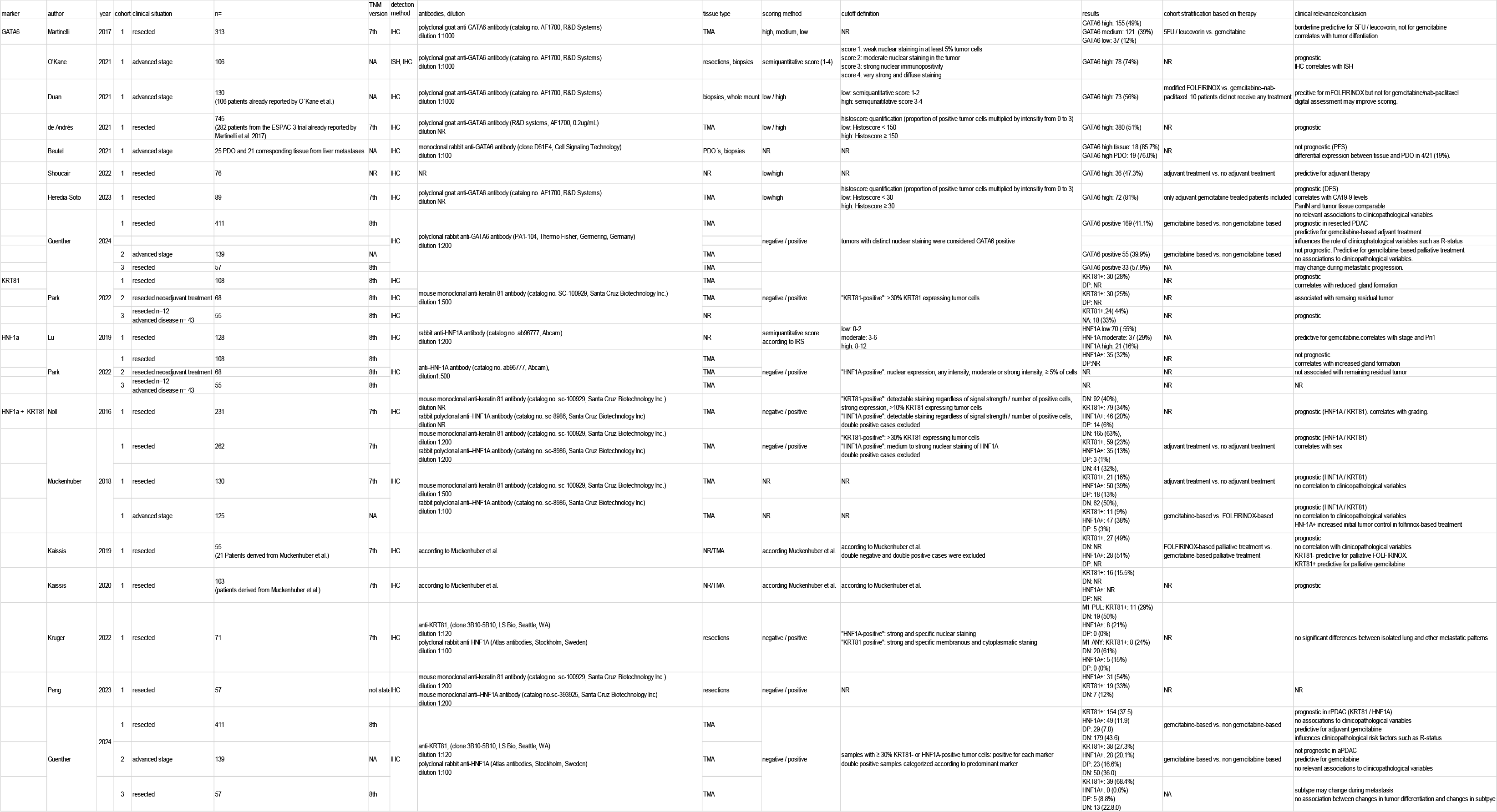
Study overview on transcriptional subtyping in PDAC.

Its retrospective nature and the fact that no patients treated with more active regimens like FFX were included in the analyses are limitations of our study. However, not only in the adjuvant setting but also in advanced disease, most patients are still treated with gemcitabine-based chemotherapy regimens to date, as co-morbidities and frailty preclude the application of potentially more efficient but more toxic regimens like FFX ^30^, which in real world data not necessarily prove to be superior to gemcitabine-based regimens ^31,32^. Thus, in the present study we propose a simple and robust diagnostic tool for therapeutic decision making in routine clinical practice. Physicians should be aware of a potential subtype switch during disease progression, which justifies a re-biopsy of metastatic lesions.

## ADDITIONAL INFORMATION

### Ethics approval and consent to participate

Each patient gave written informed consent for the use of its tumor material and clinical data for academic research purposes upon study enrollment. Ethical approval by the local ethics committee was given for each study ^33,34^ and they were carried out according to the Declaration of Helsinki. The non-commercial academic use of anonymized patient data and corresponding tumor material was approved by the ethics committee of the medical faculty of Ludwig-Maximilians-University without obtaining the patients’ informed consent (project numbers 554-11, 401-15, 20-081, 23-0222 and 23-0224).

### Availability of data and materials

The expression datasets used for validation are publicly accessible on https://portal.gdc.cancer.gov/ and https://dcc.icgc.org/.

Raw data on the patient cohorts employed in this study can be obtained from the corresponding author upon reasonable request.

### Competing interests

All authors declare no conflict of interest related to the present study.

### Funding

This study did not receive any funding.

### Author contributions

MG and SO read and scored immunohistochemical stainings, performed statistical analyses, drafted figures and wrote the manuscript. SAS analyzed bioinformatic data and drafted figures. VH, MH and SB provided clinical patient data and /or patient material. All authors read and reviewed the manuscript.

## Data Availability

The expression datasets used for validation are publicly accessible on https://portal.gdc.cancer.gov/ and https://dcc.icgc.org/. Raw data on the patient cohorts employed in this study can be obtained from the corresponding author upon reasonable request.

## Acknowledgments

We thank Andrea Sendelhofert and Anja Heier for excellent technical assistance and all study participants and their families for supporting our research.

## Supplemental figure legends

Figure S1 *Expression of KRT81, HNF1A and GATA6 in non-neoplastic tissues* KRT81 expression in normal human tonsil (A), HNF1A expression in normal human small bowel mucosa (B) and GATA6 expression in normal human non-neoplastic pancreatic ducts (C). 200-fold magnification. Scale bars indicate 50 µm.

Figure S2 Transcriptional subtypes are associated with prognosis in advanced pancreatic cancer patients Univariate analyses (Kaplan–Meier curves and log-rank tests) for PFS and OS in the subtypes based on KRT81 / HNF1A expression (A, B) as well as in the subtypes based on GATA6 expression (C, D). Crossed lines indicate censored cases.

Figure S2 Predominance of the transcriptional subtype affects prognosis in resected pancreatic cancer patients Univariate analyses (Kaplan–Meier curves and log-rank tests) for OS and DFS in the subtypes based on KRT81 and HNF1A (a, b)

Figure S3 Transcriptional subtypes are associated with prognosis in resected pancreatic cancer patients Univariate analyses (Kaplan–Meier curves and log-rank tests) for PFS and OS in the subtypes based on KRT81 / HNF1A expression (A, B) as well as in the subtypes based on GATA6 expression (C, D). Crossed lines indicate censored cases.

Figure S4 Transcriptional subtype is associated with therapy response in resected pancreatic cancer patients treated with adjuvant gemcitabine-based chemotherapy Univariate analyses (Kaplan–Meier curves and log-rank tests) for DFS and OS stratified by gemcitabine-based adjuvant treatment of the subtypes based on KRT81 / HNF1A - expression and GATA6 - expression in the TCGA-PAAD cohort (A-D) and in the PACA-CA cohort (E-H). Crossed lines indicate censored cases.

Figure S5 Transcriptional subtypes show differential expression of gemcitabine-resistance promoting genes Spearman correlations (scatter plots) of log2 mRNA expression between GATA6, HNF1A and KRT81 with ABCC3 or MVP across the TCGA-PAAD firehose dataset (A - F) as well as the ICGC-CA firehose dataset (G - L, *P < 0.05; **P < 0.01,***P < 0.0001; n.s., not significant; Spearman (two-tailed)).

Figure S6 KRT81 / HNF1A- or GATA6-based transcriptional subtypes are not associated with specific molecular alterations in pancreatic cancer Oncoprint for the ten most common genetic alterations in the subtypes based on KRT81 / HNF1A - expression (A) and GATA6 - expression (B).

## Supplemental table titles

Table S1

Prognostic impact of the predominant transcriptional subtype in advanced and resected PDAC patients.

Table S2

Comparison of both subtyping systems in the advanced and the resected PDAC cohorts.

Table S3

Assessment of transcriptional subtypes in different tissue samples for KRT81 / HNF1A (A) and GATA6 (B).

Table S4

Subtype changes during disease progression.

Table S5

Comparison of primary tumor subtype with metastatic occurrence and subtype changes for KRT81 / HNF1A and GATA6.

Table S6

Transcriptional subtypes in primary tumors and corresponding metastasis.

Table S7

Metastatic localization, primary tumor subtype and occurence of subtype change.

Table S8

Multivariate Cox regression analysis of PFS- and OS-associated factors in the advanced pancreatic cancer study cohort.

Table S9

Multivariate Cox regression analysis of PFS- and OS-associated factors in the aPDAC cohort stratified for subtypes.

Table S10

Multivariate Cox regression analysis of DFS- and OS-associated factors in the resected PDAC cohort.

Table S11

Five-year survival rates in the rPDAC cohort for subtypes based on KRT81 / HNF1A- and GATA6- expression according to adjuvant gemcitabine treatment.

Table S12

Multivariate Cox regression analysis of DFS- and OS-associated factors in the rPDAC cohort stratified for transcriptional subtypes.

## Notes

### Competing Interest Statement

The authors have declared no competing interest.

### Author Declarations

Each patient gave written informed consent for the use of its tumor material and clinical data for academic research purposes upon study enrollment. Ethical approval by the local ethics committee was given for each study 33,34 and they were carried out according to the Declaration of Helsinki. The non-commercial academic use of anonymized patient data and corresponding tumor material was approved by the ethics committee of the medical faculty of Ludwig-Maximilians-University without obtaining the patients' informed consent (project numbers 554-11, 401-15, 20-081, 23-0222 and 23-0224).

