## Supplements for "The prognostic, predictive and clinicopathological impact of KRT81 / HNF1A- and GATA6- based transcriptional subtyping in pancreatic cancer"

### Figure S1

A B C

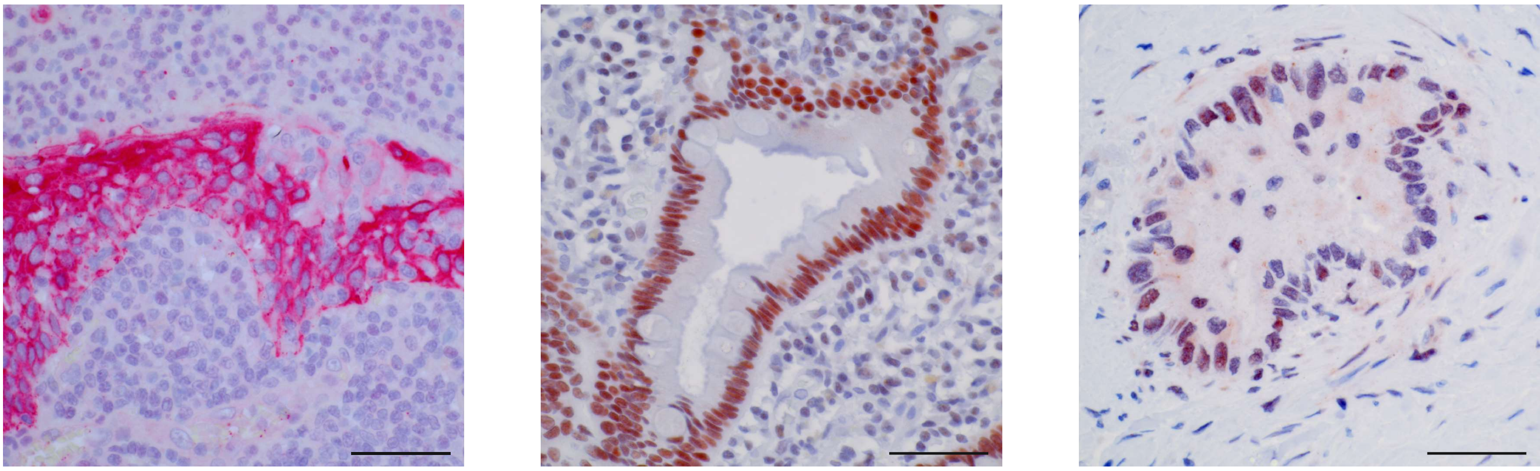

### Figure S2

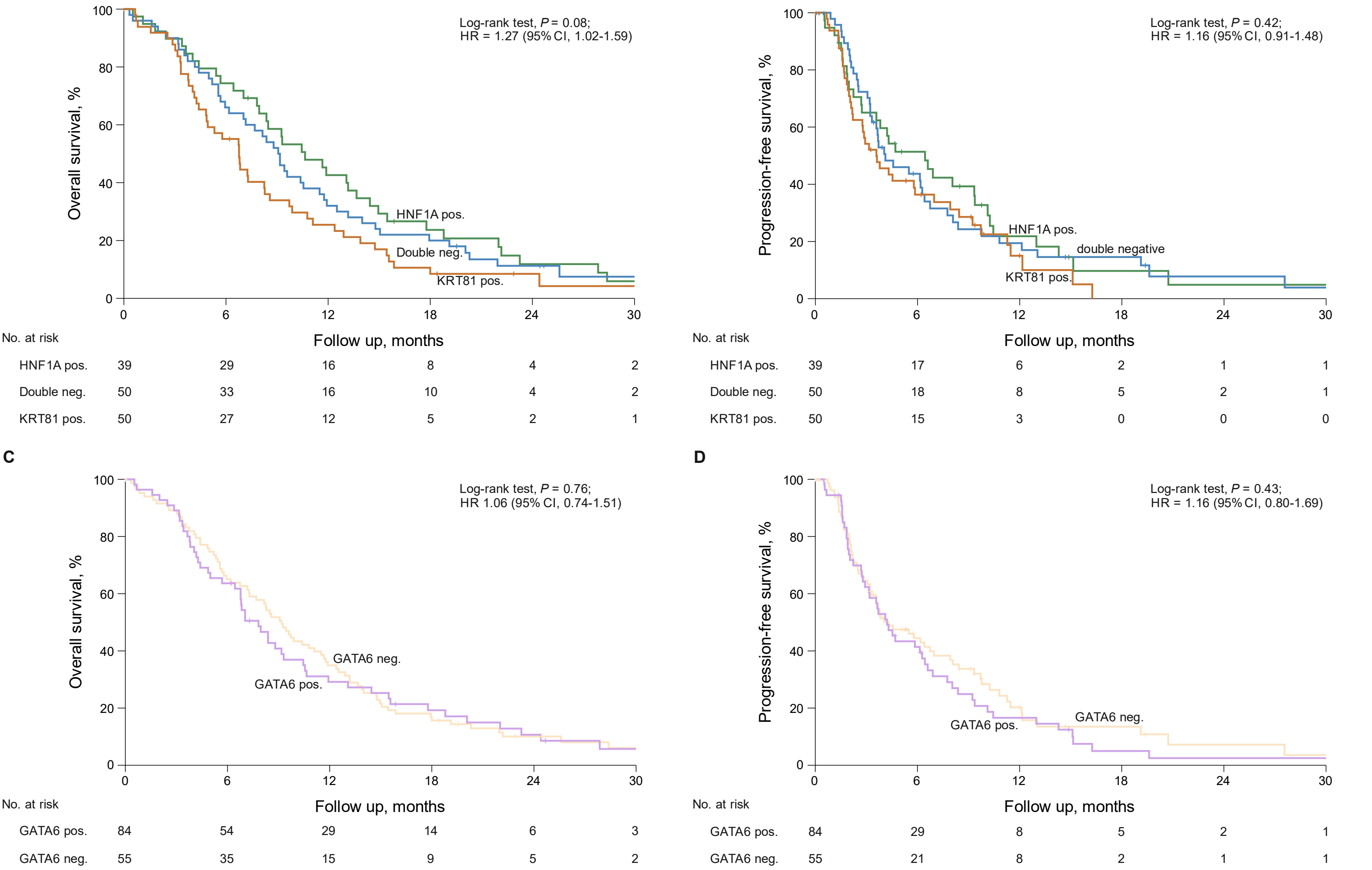

### Figure S3

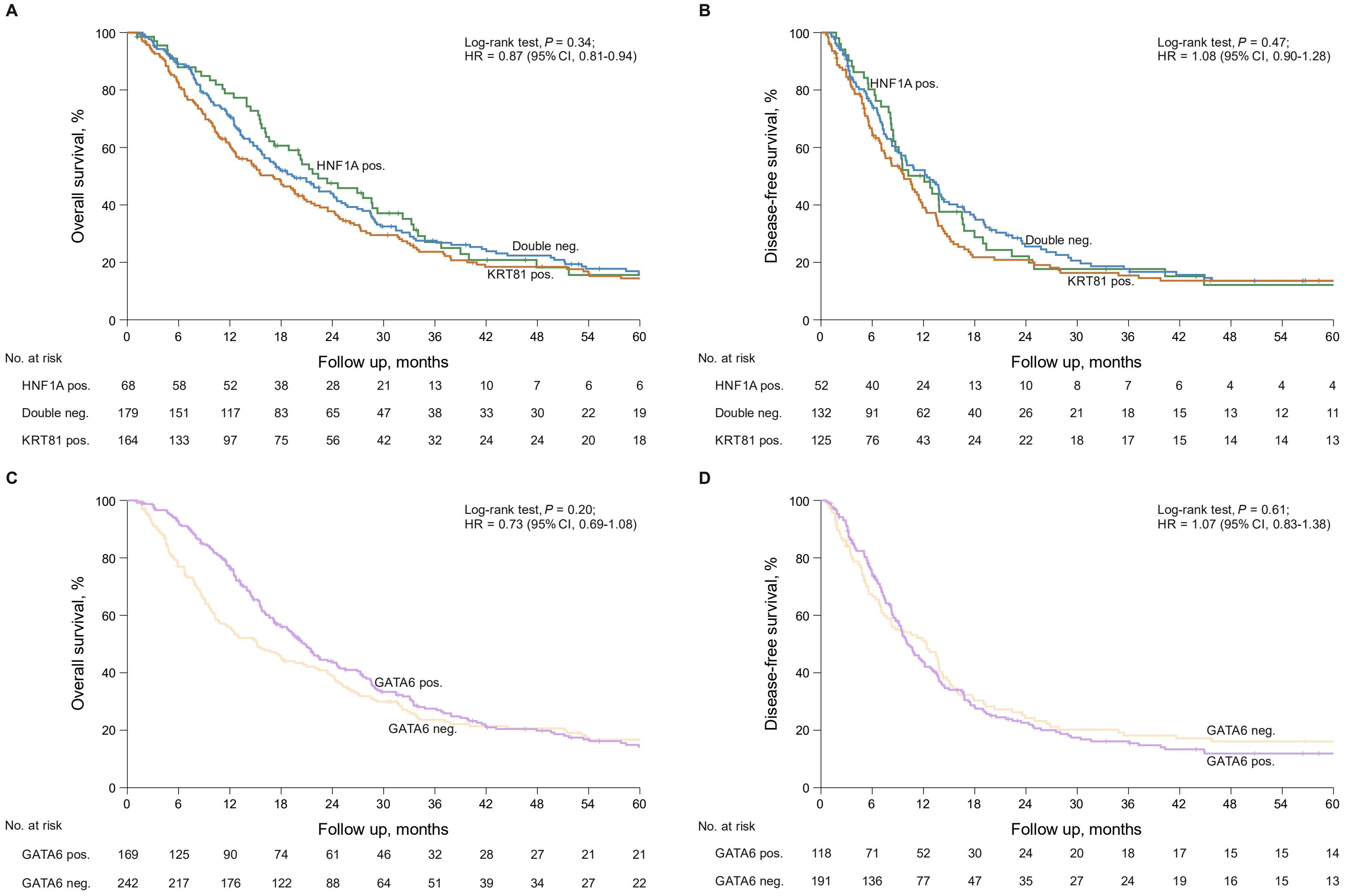
A
Log-rank test, P = 0.34;
HR = 0.87 (95% CI, 0.81-0.94)
Overall survival, %
100
80
60
40
20
0
HNF1A pos.
Double neg.
KRT81 pos.
0 6 12 18 24 30 36 42 48 54 60
Follow up, months
No. at risk
HNF1A pos. 68 58 52 38 28 21 13 10 7 6 6
Double neg. 179 151 117 83 65 47 38 33 30 22 19
KRT81 pos. 164 133 97 75 56 42 32 24 24 20 18
B
Log-rank test, P = 0.47;
HR = 1.08 (95% CI, 0.90-1.28)
Disease-free survival, %
100
80
60
40
20
0
HNF1A pos.
Double neg.
KRT81 pos.
0 6 12 18 24 30 36 42 48 54 60
Follow up, months
No. at risk
HNF1A pos. 52 40 24 13 10 8 7 6 4 4 4
Double neg. 132 91 62 40 26 21 18 15 13 12 11
KRT81 pos. 125 76 43 24 22 18 17 15 14 14 13
C
Log-rank test, P = 0.20;
HR = 0.73 (95% CI, 0.69-1.08)
Overall survival, %
100
80
60
40
20
0
GATA6 pos.
GATA6 neg.
0 6 12 18 24 30 36 42 48 54 60
Follow up, months
No. at risk
GATA6 pos. 169 125 90 74 61 46 32 28 27 21 21
GATA6 neg. 242 217 176 122 88 64 51 39 34 27 22
D
Log-rank test, P = 0.61;
HR = 1.07 (95% CI, 0.83-1.38)
Disease-free survival, %
100
80
60
40
20
0
GATA6 neg.
GATA6 pos.
0 6 12 18 24 30 36 42 48 54 60
Follow up, months
No. at risk
GATA6 pos. 118 71 52 30 24 20 18 17 15 15 14
GATA6 neg. 191 136 77 47 35 27 24 19 16 15 13

### Figure S4

A

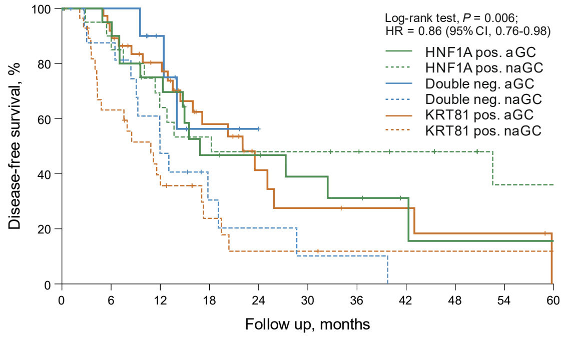

| No. at risk | 0 | 6 | 12 | 18 | 24 | 30 | 36 | 42 | 48 | 54 | 60 |
|---|---|---|---|---|---|---|---|---|---|---|---|
| HNF1A pos. aGC | 20 | 19 | 14 | 8 | 7 | 5 | 4 | 2 | 1 | 1 | 1 |
| HNF1A pos. naGC | 22 | 18 | 12 | 10 | 9 | 9 | 8 | 6 | 5 | 3 | 3 |
| Double neg. aGC | 11 | 9 | 6 | 2 | 0 | 0 | 0 | 0 | 0 | 0 | 0 |
| Double neg. naGC | 18 | 14 | 7 | 3 | 2 | 1 | 1 | 0 | 0 | 0 | 0 |
| KRT81 pos. aGC | 37 | 34 | 25 | 13 | 6 | 4 | 3 | 3 | 2 | 2 | 1 |
| KRT81 pos. naGC | 30 | 17 | 10 | 4 | 2 | 2 | 1 | 1 | 1 | 1 | 1 |

B

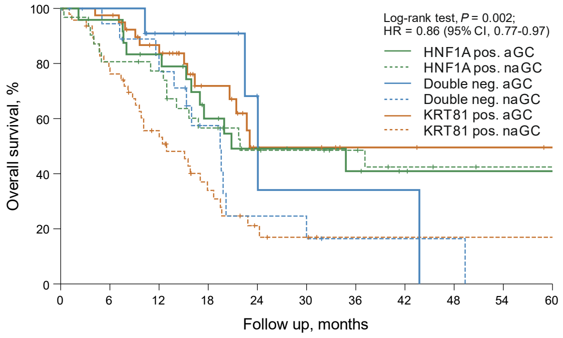

| No. at risk | 0 | 6 | 12 | 18 | 24 | 30 | 36 | 42 | 48 | 54 | 60 |
|---|---|---|---|---|---|---|---|---|---|---|---|
| HNF1A pos. aGC | 24 | 23 | 19 | 12 | 8 | 6 | 5 | 3 | 2 | 2 | 2 |
| HNF1A pos. naGC | 33 | 25 | 23 | 16 | 12 | 11 | 9 | 6 | 5 | 4 | 4 |
| Double neg. aGC | 12 | 11 | 7 | 5 | 2 | 1 | 1 | 1 | 0 | 0 | 0 |
| Double neg. naGC | 20 | 17 | 13 | 7 | 3 | 2 | 1 | 1 | 1 | 0 | 0 |
| KRT81 pos. aGC | 40 | 39 | 29 | 15 | 6 | 6 | 3 | 3 | 2 | 2 | 1 |
| KRT81 pos. naGC | 49 | 36 | 23 | 11 | 5 | 3 | 1 | 1 | 1 | 1 | 1 |

C

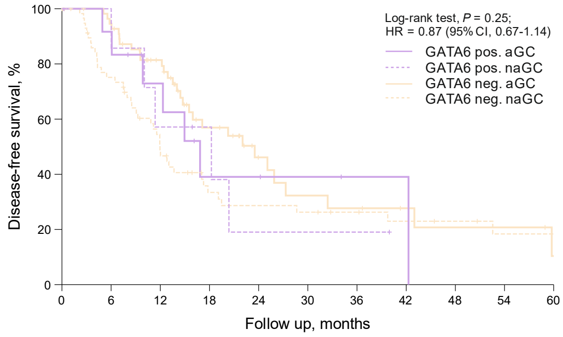

| No. at risk | 0 | 6 | 12 | 18 | 24 | 30 | 36 | 42 | 48 | 54 | 60 |
|---|---|---|---|---|---|---|---|---|---|---|---|
| Gata6 pos. aGC | 12 | 11 | 7 | 3 | 3 | 2 | 1 | 1 | 0 | 0 | 0 |
| Gata6 pos. naGC | 9 | 7 | 4 | 3 | 1 | 1 | 1 | 0 | 0 | 0 | 0 |
| Gata6 neg. aGC | 56 | 52 | 38 | 20 | 10 | 7 | 6 | 4 | 3 | 3 | 1 |
| Gata6 neg. naGC | 61 | 42 | 25 | 14 | 12 | 11 | 9 | 7 | 6 | 4 | 4 |

D

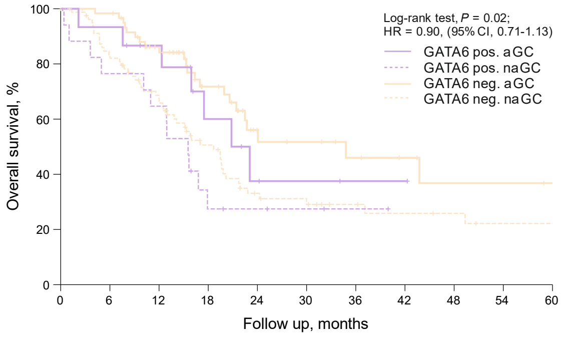

| No. at risk | 0 | 6 | 12 | 18 | 24 | 30 | 36 | 42 | 48 | 54 | 60 |
|---|---|---|---|---|---|---|---|---|---|---|---|
| Gata6 pos aGC | 15 | 14 | 11 | 6 | 3 | 2 | 1 | 1 | 0 | 0 | 0 |
| Gata6 pos.naGC | 19 | 13 | 11 | 4 | 3 | 2 | 1 | 0 | 0 | 0 | 0 |
| Gata6 neg. aGC | 61 | 59 | 44 | 26 | 13 | 11 | 8 | 6 | 4 | 4 | 3 |
| Gata6 neg. naGC | 83 | 65 | 48 | 30 | 17 | 14 | 10 | 8 | 7 | 5 | 5 |

E

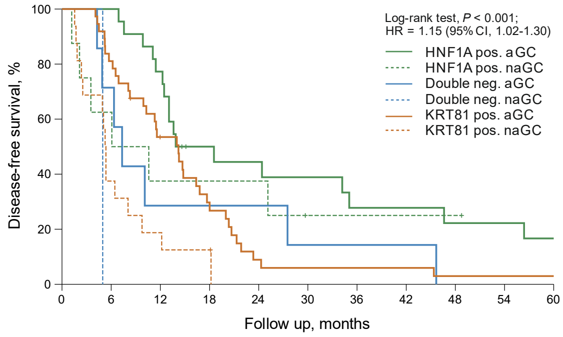

| No. at risk | 0 | 6 | 12 | 18 | 24 | 30 | 36 | 42 | 48 | 54 | 60 |
|---|---|---|---|---|---|---|---|---|---|---|---|
| HNF1A pos. aGC | 22 | 22 | 17 | 9 | 8 | 7 | 5 | 5 | 4 | 4 | 3 |
| HNF1A pos. naGC | 8 | 5 | 3 | 3 | 3 | 1 | 1 | 1 | 1 | 0 | 0 |
| Double neg. aGC | 7 | 5 | 2 | 2 | 2 | 1 | 1 | 1 | 0 | 0 | 0 |
| Double neg. naGC | 1 | 0 | 0 | 0 | 0 | 0 | 0 | 0 | 0 | 0 | 0 |
| KRT81 pos. aGC | 37 | 30 | 18 | 10 | 3 | 2 | 2 | 2 | 1 | 1 | 1 |
| KRT81 pos. naGC | 16 | 6 | 3 | 2 | 0 | 0 | 0 | 0 | 0 | 0 | 0 |

F

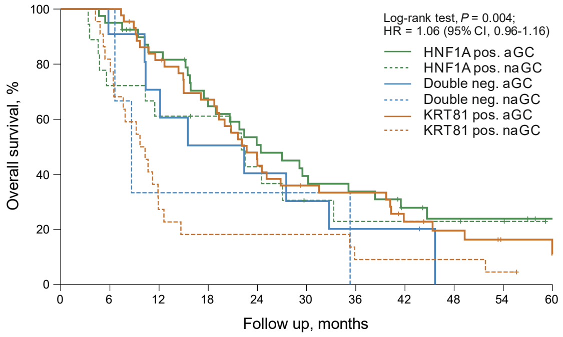

| No. at risk | 0 | 6 | 12 | 18 | 24 | 30 | 36 | 42 | 48 | 54 | 60 |
|---|---|---|---|---|---|---|---|---|---|---|---|
| HNF1A pos. aGC | 40 | 38 | 31 | 23 | 18 | 14 | 12 | 8 | 6 | 6 | 4 |
| HNF1A pos. naGC | 18 | 13 | 11 | 10 | 7 | 4 | 3 | 3 | 3 | 2 | 0 |
| Double neg. aGC | 11 | 10 | 7 | 5 | 4 | 3 | 2 | 2 | 0 | 0 | 0 |
| Double neg. naGC | 3 | 3 | 1 | 1 | 1 | 1 | 0 | 0 | 0 | 0 | 0 |
| KRT81 pos. aGC | 44 | 44 | 35 | 28 | 19 | 14 | 13 | 8 | 6 | 3 | 2 |
| KRT81 pos. naGC | 22 | 18 | 6 | 4 | 4 | 4 | 2 | 2 | 2 | 1 | 0 |

G

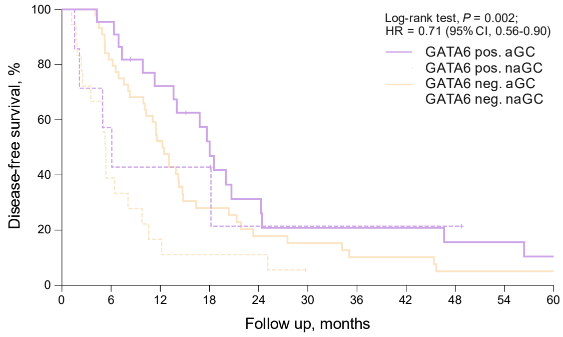

| No. at risk | 0 | 6 | 12 | 18 | 24 | 30 | 36 | 42 | 48 | 54 | 60 |
|---|---|---|---|---|---|---|---|---|---|---|---|
| Gata6 pos. aGC | 22 | 21 | 15 | 10 | 6 | 4 | 4 | 4 | 3 | 3 | 2 |
| Gata6 pos. naGC | 7 | 4 | 3 | 3 | 1 | 1 | 1 | 1 | 1 | 0 | 0 |
| Gata6 neg. aGC | 44 | 36 | 22 | 11 | 7 | 6 | 4 | 4 | 2 | 2 | 2 |
| Gata6 neg. naGC | 18 | 7 | 3 | 2 | 2 | 0 | 0 | 0 | 0 | 0 | 0 |

H

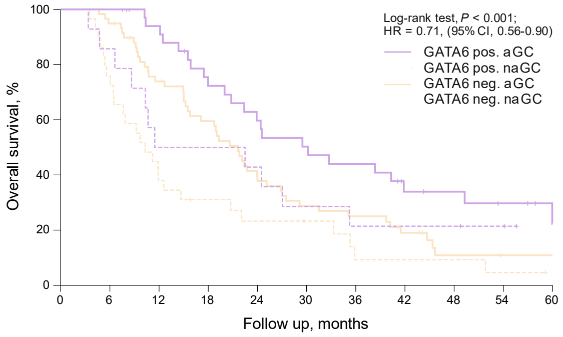

| No. at risk | 0 | 6 | 12 | 18 | 24 | 30 | 36 | 42 | 48 | 54 | 60 |
|---|---|---|---|---|---|---|---|---|---|---|---|
| Gata6 pos aGC | 36 | 36 | 31 | 23 | 19 | 16 | 14 | 9 | 8 | 6 | 3 |
| Gata6 pos.naGC | 14 | 12 | 7 | 7 | 6 | 4 | 3 | 3 | 3 | 2 | 0 |
| Gata6 neg. aGC | 59 | 56 | 42 | 33 | 22 | 15 | 13 | 9 | 4 | 3 | 3 |
| Gata6 neg. naGC | 29 | 22 | 11 | 8 | 6 | 5 | 2 | 2 | 2 | 1 | 0 |

### Figure S5

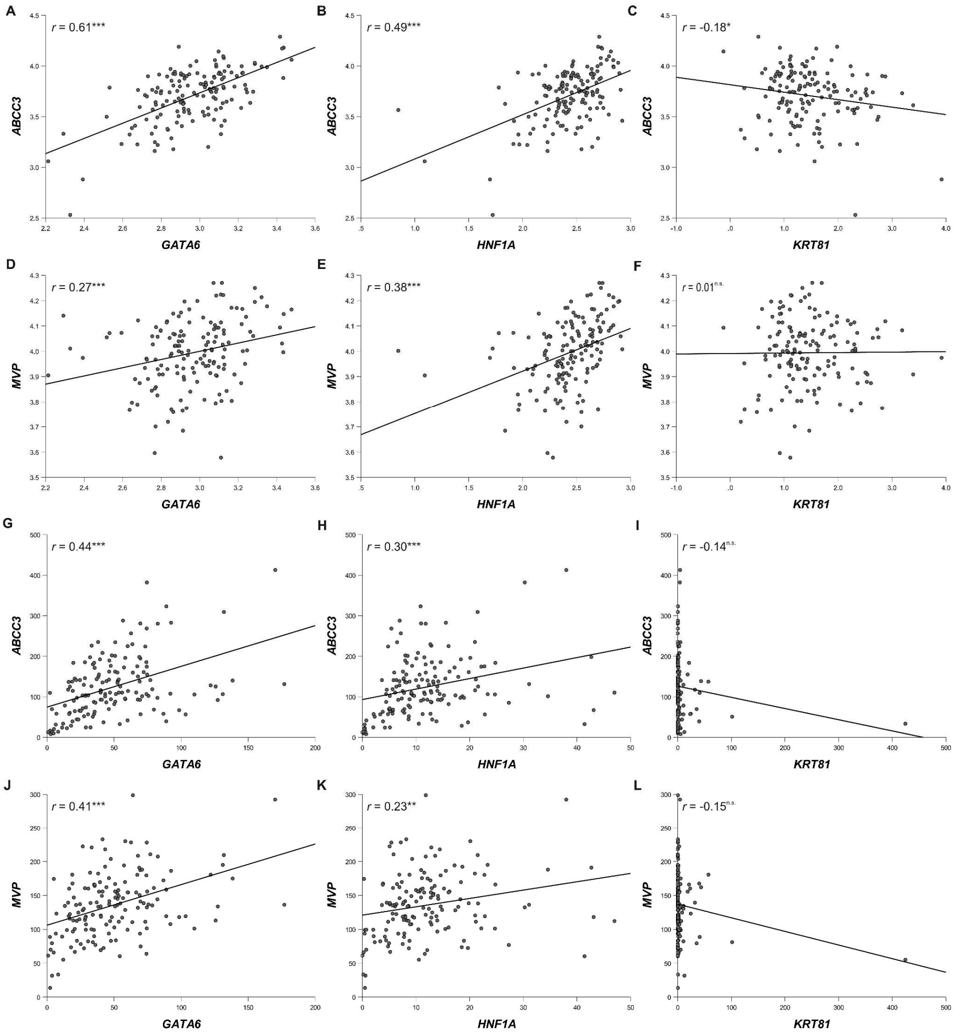

### Figure S6

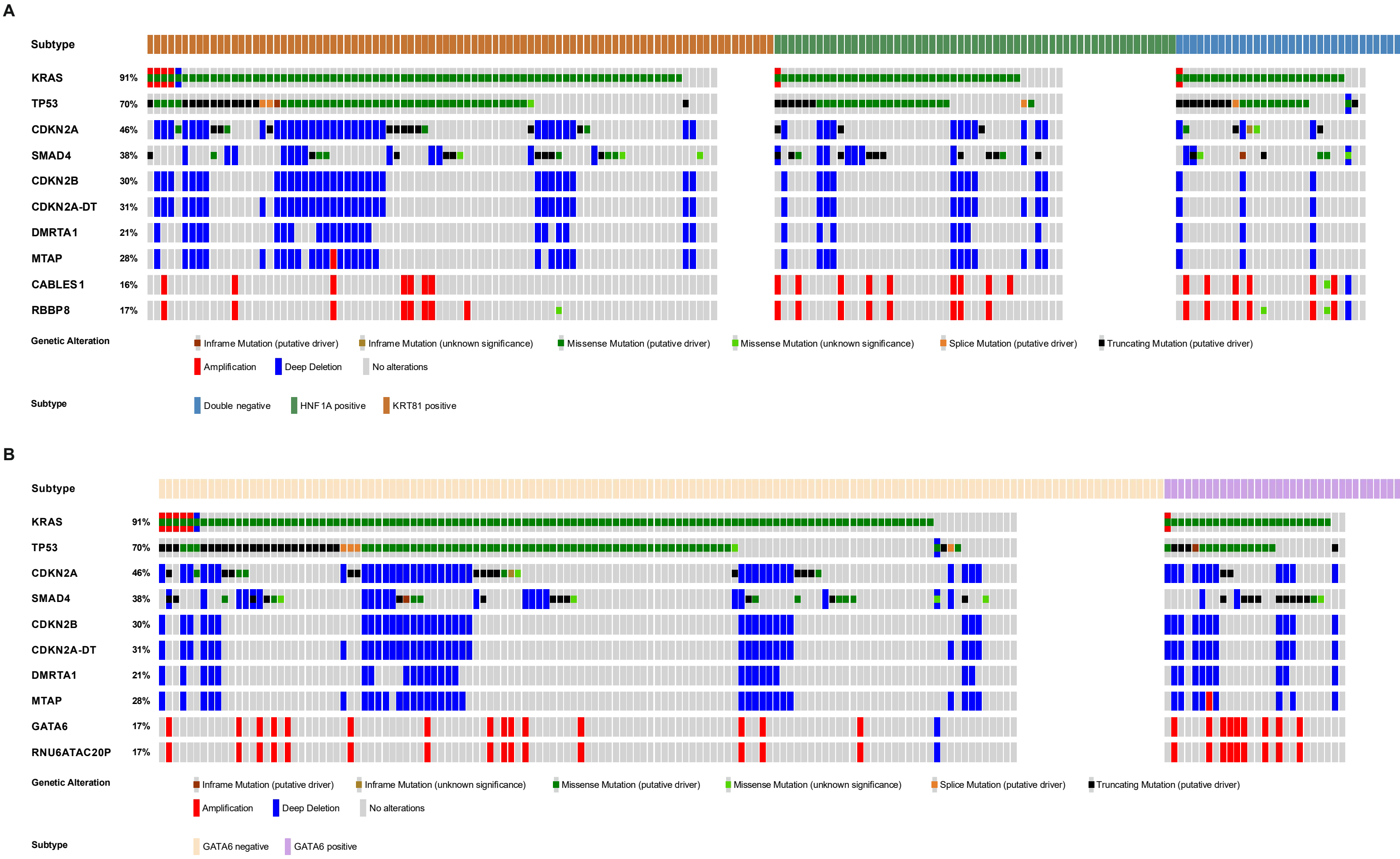

#### Table S1

Prognostic impact of the predominant transcriptional subtype in advanced and resected PDAC patients

| | advanced PDAC cohort | | | | | | | | | | resected PDAC cohort | | | | | | | | | | | |
|---|---|---|---|---|---|---|---|---|---|---|---|---|---|---|---|---|---|---|---|---|---|---|
| subtype | n | % | OS (months) | p (log-rank) | HR | 95%CI | PFS (months) | p-value (log-rank) | HR | 95%CI | n | % | OS (months) | p (log-rank) | HR | 95%CI | n | % | DFS (months) | p-value (log-rank) | HR | 95%CI |
| HNF1A pos. | 28 | 20.1 | 8.4 | 0.06 | 1.13 | 0.97 - 1.31 | 3.8 | 0.01 | 1.05 | 0.89 - 1.23 | 49 | 11.9 | 22.3 | 0.49 | 1.07 | 0.95 - 1.19 | 35 | 11.3 | 9.5 | 0.76 | 1.04 | 0.92 - 1.18 |
| HNF1A pos. (double pos.) | 11 | 7.9 | 17.8 | | | | 15.2 | | | | 19 | 4.6 | 34.8 | | | | 17 | 5.5 | 13.8 | | | |
| double neg. | 50 | 36.0 | 9.1 | | | | 4.1 | | | | 179 | 43.6 | 19.3 | | | | 132 | 42.7 | 12.4 | | | |
| KRT81 pos. | 38 | 27.3 | 6.8 | | | | 3.6 | | | | 154 | 37.5 | 17.1 | | | | 116 | 37.5 | 9.5 | | | |
| KRT81 pos. (double pos.) | 12 | 8.6 | 6.8 | | | | 2.9 | | | | 10 | 2.4 | 15.5 | | | | 9 | 2.9 | 16.8 | | | |

#### Table S2

Comparison of both subtyping systems in the advanced and resected PDAC cohort

| | aPDAC cohort | | | rPDAC cohort | | |
|---|---|---|---|---|---|---|
| | GATA6 neg. | GATA6 pos. | p-value (χ2-test) | GATA6 neg. | GATA6 pos. | p-value (χ2-test) |
| HNF1A pos. | 14 (16.7) | 25 (45.5) | <0.001 | 7 (4.1) | 61 (25.2) | < 0.001 |
| double neg. | 37 (44.0) | 13 (23.6) | | 66 (39.1) | 113 (46.7) | |
| KRT81 pos. | 33 (39.3) | 17 (30.9) | | 96 (56.8) | 68 (28.1) | |

#### Table S3

Assessment of transcriptional subtypes in different tissue samples for KRT81 / HNF1A (A) and GATA6 (B)

A

| | | subtype repunched | | | |
|---|---|---|---|---|---|
| | | HNF1a pos. | double negative | KRT81 pos. | p-value (χ2-test) |
| subtype resected cohort | HNF1a pos. | 2 (100.0) | 0 (0.0) | 1 (7.7) | <0.001 |
| | double negative | 0 (0.0) | 8 (80.0) | 4 (30.8) | |
| | KRT81 pos. | 0 (0.0) | 2 (20.0) | 8 (61.5) | |

B

| | | subtype repunched | | |
|---|---|---|---|---|
| | | GATA6 negative | GATA6 positive | p-value (χ2-test) |
| subtype resected cohort | GATA6 negative | 7 (87.%) | 1 (5.9) | <0.001 |
| | GATA6 positive | 1 (12.5) | 16 (94.1) | |

#### Table S4

Subtype changes during disease progression

| | KRT81/HNF1A | | | GATA6 | | |
|---|---|---|---|---|---|---|
| | subtype switch | | | subtype switch | | |
| metastasis type | no | yes | p-value (χ2-test) | no | yes | p-value (χ2-test) |
| synchronous | 25 (64.1) | 8 (44.4) | 0.16 | 20 (48.8) | 13 (81.3) | 0.03 |
| metachronous | 14 (35.9) | 10 (55.6) | | 21 (51.2) | 2 (18.8) | |

#### Table S5

Comparison of primary tumor subtype with metastatic occurrence and subtype changes for KRT81 / HNF1A and GATA6

| | subtype primary tumor, no (%) | | | | subtype primary tumor, no (%) | | |
|---|---|---|---|---|---|---|---|
| | KRT81 pos. (n=37) | double neg. (n=17) | HNF1A pos. (n=3) | p-value (χ2-test ) | GATA6 neg. (n=23) | GATA6 pos. (n=34) | p-value (χ2-test ) |
| subtype change | | | | | | | |
| no switch | 25 (67.6) | 13 (76.5) | 1 (33.3) | 0.33 | 16 (69.6) | 25 (73.5) | 0.74 |
| switch | 12 (32.4) | 4 (23.5) | 2 (66.7) | | 7 (30.4) | 9 (26.5) | |
| metastasis type | | | | | | | |
| synchronous | 23 (62.2) | 9 (52.9) | 1 (33.3) | 0.55 | 13 (56.5) | 20 (58.8) | 0.86 |
| metachronous | 14 (37.8) | 8 (47.1) | 2 (66.7) | | 10 (43.5) | 14 (41.2) | |
| prognostic properties of subtype switch | | | | | | | |
| subtype constant | 25 (68.4) | 13 (76.5) | 1 (33.3) | <0.001 | 16 (69.6) | 25 (73.5) | <0.001 |
| prognostically more favorable | 12 (32.4) | 3 (17.6) | 0 (0.0) | | 7 (30.4) | 0 (0.0) | |
| prognostically less favorable | 0 (0.0) | 1 (5.9) | 2 (66.7) | | 0 (0.0) | 9 (26.5) | |

#### Table S6

Transcriptional subtypes in primary tumors and corresponding metastasis

| | | subtype metastasis | | | |
|---|---|---|---|---|---|
| | | HNF1a pos. | double neg. | KRT81 pos. | p-value (χ2-test) |
| subtype primary tumor | HNF1a pos. | 1 (16.7) | 0 (0.0) | 2 (7.1) | <0.001 |
| | double neg. | 3 (50.0) | 13 (56.5) | 1 (3.6) | |
| | KRT81 pos. | 2 (33.3) | 10 (43.5) | 25 (89.3) | |
| | | GATA6 pos. | GATA6 neg. | p-value (χ2-test) | |
| | GATA6 pos. | 16 (64.0) | 7 (21.9) | 0.001 | |
| | GATA6 neg. | 9 (36.0) | 25 (78.1) | | |

#### Table S7

Metastatic localization, primary tumor subtype and occurence of subtype change

| | metastasis localization | | | | | |
|---|---|---|---|---|---|---|
| | HEP | PER | PUL | PLE | OTH | p-value (χ2-test) |
| subtype primary tumor | | | | | | |
| HNF1A positive | 0 (0.0) | 2 (12.5) | 0 (0.0) | 1 (50.0) | 0 (0.0) | 0.10 |
| double negative | 8 (26.7) | 5 (31.3) | 2 (50.0) | 0 (0.0) | 2 (40.0) | |
| KRT81 positive | 22 (73.3) | 9 (56.3) | 2 (50.0) | 1 (50.0) | 3 (80.0) | |
| subtype metastasis | | | | | | |
| HNF1A positive | 2 (6.7) | 3 (18.8) | 1 (25.0) | 0 (0.0) | 0 (0.0) | 0.28 |
| double negative | 13 (43.3) | 4 (25.0) | 3 (75.0) | 0 (0.0) | 3 (60.0) | |
| KRT81 positive | 15 (50.0) | 9 (56.3) | 0 (0.0) | 2 (100.0) | 2 (40.0) | |
| subtype switch (KRT81 / HNF1A) | | | | | | |
| no switch | 21 (70.0) | 11 (68.8) | 2 (50.0) | 1 (50.0) | 4 (80.0) | 0.86 |
| switch | 9 (30.0) | 5 (31.1) | 2 (50.0) | 1 (50.0) | 1 (20.0) | |
| subtype primary tumor | | | | | | |
| GATA6 negative | 10 (33.3) | 10 (62.5) | 2 (50.0) | 0 (0.0) | 1 (20.0) | 0.18 |
| GATA6 positive | 20 (66.7) | 6 (37.5) | 2 (50.0) | 2 (100.0) | 4 (80.0) | |
| subtype metastasis | | | | | | |
| GATA6 negative | 16 (53.3) | 7 (43.8) | 1 (25.0) | 0 (0.0) | 1 (20.0) | 0.36 |
| GATA6 positive | 14 (46.7) | 9 (56.3) | 3 (75.0) | 2 (100.0) | 4 (80.0) | |
| subtype switch (GATA6) | | | | | | |
| no switch | 22 (73.3) | 11 (68.8) | 3 (75.0) | 2 (100.0) | 3 (60.0) | 0.87 |
| switch | 8 (26.7) | 5 (31.3) | 1 (25.0) | 0 (0.0) | 2 (40.0) | |

#### Table S8

Multivariate Cox regression analysis of PFS- and OS-associated factors in the aPDAC cohort.

| | OS | | | |
|---|---|---|---|---|
| | parameter | p-value (Cox) | HR | 95%CI |
| KRT81 / HNF1A | HNF1A pos. | 0.07 | | |
| | double neg. | 1.00 | 1.00 | 0.63 - 1.55 |
| | KRT81 pos. | 0.06 | 1.55 | 1.01 - 2.50 |
| | grade group | 0.009 | 1.64 | 1.13 - 2.39 |
| | CTX type | 0.09 | 1.85 | 1.12 - 3.04 |
| | disease stage at therapy initiation | 0.02 | 1.85 | 1.12 - 3.04 |
| GATA6 | grade group | 0.002 | 1.77 | 1.23 - 2.54 |
| | disease stage at therapy initiation | 0.02 | 1.81 | 1.10 - 2.98 |

| | PFS | | | |
|---|---|---|---|---|
| | parameter | p-value (Cox) | HR | 95%CI |
| KRT81 / HNF1A | grade group | 0.002 | 1.89 | 1.27 - 2.80 |
| | CTX type | < 0.001 | 0.43 | 0.28 - 0.65 |
| | disease stage at therapy initiation | 0.04 | 1.74 | 1.03 - 2.94 |
| GATA6 | grade group | 0.002 | 1.89 | 1.27 - 2.80 |
| | CTX type | < 0.001 | 0.43 | 0.28 - 0.65 |
| | disease stage at therapy initiation | 0.04 | 1.74 | 1.03 - 2.94 |

#### Table S9

Multivariate Cox regression analysis of PFS- and OS-associated factors in the aPDAC cohort stratified for subtypes.

| | PFS | | | |
|---|---|---|---|---|
| | parameter | p-value (Cox) | HR | 95%CI |
| GATA6 negative | grade group | 0.04 | 1.79 | 1.02 - 3.14 |
| | CTX type | < 0.001 | 0.33 | 0.18 - 0.59 |
| GATA6 postive | disease stage at therapy initiation | 0.04 | 2.57 | 1.04 - 6.34 |
| | grade group | 0.06 | 1.77 | 0.97 - 3.21 |
| KRT81 positive | KPS group | 0.03 | 0.45 | 0.22 - 0.93 |
| | CTX type | 0.02 | 0.41 | 0.19 - 0.87 |
| | disease stage at therapy initiation | 0.06 | 2.31 | 0.96 - 5.60 |
| double negative | CTX type | < 0.001 | 0.23 | 0.11 - 0.47 |
| | disease stage at therapy initiation | 0.07 | 2.47 | 0.94 - 6.49 |
| HNF1A positive | grade group | 0.013 | 2.59 | 1.22 - 5.49 |

| | OS | | | |
|---|---|---|---|---|
| | parameter | p-value (Cox) | HR | 95%CI |
| GATA6 negative | grade group | 0.002 | 2.17 | 1.33 - 3.54 |
| GATA6 postive | disease stage at therapy initiation | 0.10 | 2.00 | 0.89 - 4.54 |
| KRT81 positive | KPS group | 0.007 | 0.42 | 0.22 - 0.79 |
| | disease stage at therapy initiation | 0.04 | 2.40 | 1.05 - 5.51 |
| double negative | CTX type | 0.04 | 0.52 | 0.28 - 0.96 |
| HNF1A postive | grade group | 0.07 | 1.88 | 0.95 - 3.71 |

#### Table S10

Multivariate Cox regression analysis of DFS- and OS-associated factors in the resected PDAC cohort.

| | OS | | | |
|---|---|---|---|---|
| | parameter | p-value (Cox) | HR | 95%CI |
| KRT81 / HNF1A | HNF1A-positive | 0.04 | | |
| | Double-negative | 0.43 | 1.15 | 0.82 - 1.60 |
| | KRT81-positive | 0.03 | 1.46 | 1.04 - 2.05 |
| | pT1a | 0.006 | | |
| | pT1b | 0.81 | 0.84 | 0.19 - 3.67 |
| | pT1c | 0.33 | 0.54 | 0.16 - 1.85 |
| | pT2 | 0.44 | 0.62 | 0.19 - 2.06 |
| | pT3 | 0.99 | 1.00 | 0.30 - 3.32 |
| | pT4 | 0.44 | 0.55 | 0.12 - 2.51 |
| | R-status | < 0.001 | 1.51 | 1.19 - 1.91 |
| | grade group | < 0.001 | 1.64 | 1.27 - 2.12 |
| | pN0 | < 0.001 | | |
| | pN1 | 0.14 | 1.22 | 0.94 - 1.58 |
| | pN2 | < 0.001 | 2.12 | 1.59 - 2.83 |
| | adjvant gemcitabine treatment | < 0.001 | 0.47 | 0.38 - 0.60 |
| GATA6 | GATA6-positive | 0.006 | 0.73 | 0.58 - 0.91 |
| | pT1a | 0.006 | | |
| | pT1b | 0.88 | 0.90 | 0.21 - 3.81 |
| | pT1c | 0.33 | 0.56 | 0.17 - 1.86 |
| | pT2 | 0.48 | 0.66 | 0.20 - 2.13 |
| | pT3 | 0.95 | 1.04 | 0.32 - 3.40 |
| | pT4 | 0.52 | 0.61 | 0.14 - 2.71 |
| | R-Status | 0.001 | 1.49 | 1.17 - 1.88 |
| | grade group | <0.001 | 1.65 | 1.28 - 2.13 |
| | pN0 | <0.001 | | |
| | pN1 | 0.11 | 1.24 | 0.95 - 1.60 |
| | pN2 | <0.001 | 2.15 | 1.62 - 2.87 |
| | adjvant gemcitabine treatment | <0.001 | 0.47 | 0.38 - 0.60 |

| | DFS | | | |
|---|---|---|---|---|
| | parameter | p-value (Cox) | HR | 95%CI |
| KRT81 / HNF1A | pT1a | 0.009 | | |
| | pT1b | 0.81 | 1.25 | 0.21 - 7.51 |
| | pT1c | 0.99 | 0.99 | 0.23 - 4.26 |
| | pT2 | 0.68 | 1.35 | 0.33 - 5.58 |
| | pT3 | 0.29 | 2.18 | 0.52 - 9.15 |
| | pT4 | 0.87 | 1.15 | 0.20 - 6.61 |
| | grade group | 0.02 | 1.40 | 1.07 - 1.84 |
| | pN0 | < 0.001 | | |
| | pN1 | 0.35 | 1.15 | 0.86 - 1.55 |
| | pN2 | < 0.001 | 1.94 | 1.39 - 2.72 |
| | adjvant gemcitabine treatment | < 0.001 | 0.59 | 0.45 - 0.77 |
| GATA6 | pT1a | 0.009 | | |
| | pT1b | 0.81 | 1.25 | 0.21 - 7.51 |
| | pT1c | 0.99 | 0.99 | 0.23 - 4.26 |
| | pT2 | 0.68 | 1.35 | 0.33 - 5.58 |
| | pT3 | 0.29 | 2.18 | 0.52 - 9.15 |
| | pT4 | 0.87 | 1.15 | 0.20 - 6.61 |
| | grade group | 0.02 | 1.40 | 1.07 - 1.84 |
| | pN0 | < 0.001 | | |
| | pN1 | 0.35 | 1.15 | 0.86 - 1.55 |
| | pN2 | < 0.001 | 1.94 | 1.39 - 2.72 |
| | adjvant gemcitabine treatment | < 0.001 | 0.59 | 0.45 - 0.77 |

#### Table S11

Five-year survival rates for subtypes based on KRT81 / HNF1A and GATA6 expression according to adjuvant gemcitabine treatment

| | | adjuvant gemcitabine treatment | | |
|---|---|---|---|---|
| | 5a survival rate | no | yes | p-value (χ2-test) |
| GATA6 neg. | dead | 77 (98.7) | 54 (73.0) | <0.001 |
| | alive | 1 (1.3) | 20 (27.0) | |
| GATA6 pos. | dead | 78 (90.7) | 103 (88.0) | 0.55 |
| | alive | 8 (9.3) | 14 (12.0) | |
| KRT81 pos. | dead | 74 (97.4) | 57 (78.1) | <0.001 |
| | alive | 2 (2.6) | 16 (21.9) | |
| double neg. | dead | 56 (91.8) | 75 (84.3) | 0.17 |
| | alive | 5 (8.2) | 14 (15.7) | |
| HNF1A pos. | dead | 25 (92.6) | 25 (86.2) | 0.44 |
| | alive | 2 (7.4) | 4 (13.8) | |

**Table S12**

Multivariate Cox regression analysis of DFS- (A) and OS (B)-associated factors in the rPDAC cohort stratified for transcriptional subtypes.

| | | | | |
|---|---|---|---|---|
| | DFS | | | |
| | parameter | p-value (Cox) | HR | 95%CI |
| GATA6 negative | R-status | 0.08 | 1.51 | 0.95 - 2.40 |
| | pN0 | 0.007 | | |
| | pN1 | 0.03 | 0.58 | 0.36 - 0.96 |
| | pN2 | 0.12 | 1.53 | 0.89 - 2.62 |
| | adjuvant gemcitabine treatment | <0.001 | 0.28 | 0.18 - 0.44 |
| GATA6 postive | pT1a | 0.01 | | |
| | pT1b | 0.47 | 0.55 | 0.11 - 2.75 |
| | pT1c | 0.14 | 0.39 | 0.11 - 1.34 |
| | pT2 | 0.12 | 0.39 | 0.12 - 1.28 |
| | pT3 | 0.56 | 0.70 | 0.21 - 2.34 |
| | pT4 | 0.45 | 0.54 | 0.11 - 2.63 |
| | grade group | <0.001 | 1.99 | 1.41 - 2.82 |
| | pN0 | <0.001 | | |
| | pN1 | 0.02 | 1.53 | 1.09 - 2.16 |
| | pN2 | <0.001 | 2.22 | 1.53 - 3.24 |
| KRT81 positive | UICC stage IA | 0.006 | | |
| | UICC stage IB | 0.66 | 1.25 | 0.46 - 3.38 |
| | UICC stage IIA | 0.16 | 2.26 | 0.72 - 7.14 |
| | UICC stage IIB | 0.178 | 1.93 | 0.74 - 5.03 |
| | UICC stage III | 0.02 | 3.30 | 1.18 - 9.23 |
| | UICC stage IV | 0.007 | 4.67 | 1.51 - 14.43 |
| | R-status | 0.003 | 2.02 | 1.28 - 3.20 |
| | adjuvant gemcitabine treatment | <0.001 | 0.29 | 0.19 - 0.45 |
| double negative | pT1c | 0.03 | | |
| | pT1c | 0.95 | 0.95 | 0.20 - 4.43 |
| | pT2 | 0.91 | 0.91 | 0.18 - 4.51 |
| | pT3 | 0.30 | 2.36 | 0.46 - 12.04 |
| | pT4 | 0.34 | 2.49 | 0.38 - 16.53 |
| | UICC stage IA | 0.009 | | |
| | UICC stage IB | 0.53 | 1.45 | 0.46 - 4.57 |
| | UICC stage IIA | 0.43 | 0.60 | 0.17 - 2.15 |
| | UICC stage IIB | 0.78 | 0.86 | 0.31 - 2.43 |
| | UICC stage III | 0.18 | 2.14 | 0.71 - 6.43 |
| | UICC stage IV | 0.48 | 0.67 | 0.22 - 2.05 |
| | grade group | 0.09 | 1.46 | 0.94 - 2.26 |
| HNF1A positive | pT1a | 0.03 | | |
| | pT1c | 0.86 | 1.22 | 0.14 - 10.67 |
| | pT2 | 0.63 | 1.65 | 0.22 - 12.60 |
| | pT3 | 0.14 | 4.89 | 0.61 - 39.53 |
| | pN0 | 0.02 | | |
| | pN1 | 0.02 | 2.42 | 1.14 - 5.13 |
| | pN2 | 0.02 | 2.67 | 1.19 - 5.97 |
| | OS | | | |
| | parameter | p-value (Cox) | HR | 95%CI |
| GATA6 negative | R-status | 0.003 | 1.78 | 1.22 - 2.59 |
| | pN0 | 0.02 | | |
| | pN1 | 0.46 | 0.86 | 0.58 - 1.28 |
| | pN2 | 0.03 | 1.63 | 1.05 - 2.55 |
| | adjuvant gemcitabine treatment | <0.001 | 0.24 | 0.16 - 0.35 |
| GATA6 positive | pT1a | 0,02 | | |
| | pT1b | 0,66 | 0,67 | 0.11 - 4.05 |
| | pT1c | 0,08 | 0,31 | 0.08 - 1.13 |
| | pT2 | 0,05 | 0,29 | 0.09 - 1.01 |
| | pT3 | 0,50 | 0,65 | 0.18 - 2.30 |
| | pT4 | 0,58 | 0,63 | 0.12 - 3.25 |
| | grade group | 0,001 | 2,19 | 1.38 - 3.46 |
| | pN0 | 0,02 | | |
| | pN1 | 0,30 | 1,29 | 0.80 - 2.07 |
| | pN2 | 0,006 | 2,03 | 1.23 - 3.36 |
| KRT81 positive | UICC stage IA | 0.004 | | |
| | UICC stage IB | 0.75 | 0.85 | 0.32 - 2.28 |
| | UICC stage IIA | 0.28 | 1.79 | 0.63 - 5.11 |
| | UICC stage IIB | 0.63 | 1.26 | 0.50 - 3.19 |
| | UICC stage III | 0.20 | 1.92 | 0.71 - 5.19 |
| | UICC stage IV | 0.05 | 2.91 | 1.03 - 8.23 |
| | R-status | <0.001 | 2.43 | 1.67 - 3.55 |
| | grade group | 0.02 | 1.59 | 1.06 - 2.37 |
| | adjuvant gemcitabine treatment | <0.001 | 0.24 | 0.17 - 0.35 |
| double negative | UICC stage IA | 0.01 | | |
| | UICC stage IB | 0.64 | 1.19 | 0.58 - 2.46 |
| | UICC stage IIA | 0.67 | 1.20 | 0.53 - 2.70 |
| | UICC stage IIB | 0.93 | 1.02 | 0.53 - 2.03 |
| | UICC stage III | 0.01 | 2.38 | 1.19 - 4.74 |
| | UICC stage IV | 0.85 | 1.08 | 0.50 - 2.31 |
| | R-status | 0.04 | 1.45 | 1.01 - 2.07 |
| | grade group | <0.001 | 1.97 | 1.33 - 2.91 |
| HNF1A postive | pT1a | 0.002 | | |
| | pT1c | 0.60 | 0.64 | 0.13 - 3.34 |
| | pT2 | 0.59 | 0.66 | 0.15 - 2.97 |
| | pT3 | 0.25 | 2.49 | 0.52 - 11.88 |
| | pN0 | 0.002 | | |
| | pN1 | 0.01 | 2.57 | 1.24 - 5.34 |
| | pN2 | <0.001 | 3.27 | 1.63 - 6.56 |
